# Platelet CKB expression is associated with elevated serum CK-BB and metastatic phenotype in lung cancer

**DOI:** 10.1101/2025.07.03.25330795

**Authors:** Lubei Rao, Chao Hu, Yan Yue, Yajun Luo, Bo Ye, Yue Liang, Guanbin Zhang, Dongsheng Wang

**Author notes:** Correspondence (L. R.); Department of Clinical Laboratory, Sichuan Clinical Research Center for Cancer, Sichuan Cancer Hospital & Institute, Sichuan Cancer Center, University of Electronic Science and Technology of China, Chengdu, China. No. 55, Section 4, Renmin South Road, Wuhou District, Chengdu City, Sichuan Province, China. (L. R.); (Y. Y.); (B. Y.); (D. W.); (C. H.); (Y. L.); (Y. L.); (G. Z.).

## Abstract

The abnormal increase of serum creatine kinase-MB activity exceeding total creatine kinase activity is an uncommon and paradoxical laboratory finding often observed in patients with cancer. However, the underlying origin and clinical relevance of this phenomenon remain unclear. In this study, we analyzed a large cohort of cancer patients presenting with this biochemical abnormality. We first characterized the cancer types and stages most frequently associated with this pattern, finding enrichment in colorectal and lung cancers, particularly at advanced stages. Serum isoenzyme electrophoresis revealed that the abnormal elevation was not due to myocardial damage but instead driven by high levels of brain-type and mitochondrial creatine kinase isoforms. We introduced a new composite index integrating these non-cardiac isoenzymes, which was correlated with poor prognosis, especially in colorectal cancer. Notably, we identified an unexpected elevation of creatine kinase brain-type messenger RNA in platelets from lung cancer patients. This expression level was significantly higher than that found in serum and showed a strong positive correlation, suggesting that platelets may contribute to the circulating isoenzyme pool. This observation was supported by analysis of a public dataset of tumor-educated platelets, which confirmed higher creatine kinase expression in lung cancer patients. Additional single-cell sequencing data showed that distant metastatic lesions, but not primary tumors, exhibited elevated creatine kinase expression. Functional gene enrichment analysis revealed associated pathways involved in metabolism, oxidative stress, and intercellular signaling. These findings suggest that platelet-related creatine kinase may be linked to metastatic potential in lung cancer and represent a previously underrecognized component in cancer-associated biochemical alterations.

## 1. Introduction

Creatine kinase (CK) is a central regulator of cellular energy homeostasis (1). It catalyzes the reversible transfer of a phosphate group from adenosine triphosphate (ATP) to creatine, forming phosphocreatine and adenosine diphosphate (ADP) (2). Through this reaction, CK plays a vital role in tissues with high energy demands, such as skeletal muscle, heart, and brain. CK is encoded by multiple genes, forming tissue-specific isoenzyme subtypes. Among them, CKB encodes the creatine kinase brain-type isoenzyme (CK-BB); CKM encodes the creatine kinase muscle-type isoenzyme (CK-MM); and the hybrid creatine kinase-MB isoenzyme (CK-MB) is a heterodimer composed of CKB and CKM gene products. These isoforms are predominantly cytoplasmic (3, 4). In addition, mitochondrial creatine kinase (MtCK), encoded by the CKMT1 and CKMT2 genes, is predominantly located within mitochondria, and exists in the form of mitochondrial creatine kinase isoenzyme II (CKII, dimeric form) and mitochondrial creatine kinase isoenzyme I (CKI, monomeric form) (5, 6), playing a key role in ATP buffering in oxidative tissues.

In healthy individuals, the serum CK pool is predominantly composed of CK-MM, with negligible amounts of other isoenzymes. During myocardial injury, CK-MB is released into the circulation, and serum CK-MB activity measurement is widely used for the diagnosis of myocardial infarction (7–11). However, serum CK-MB activity detected by the immunoinhibition assay (12) may be influenced by the presence of non-CK-MB isoenzymes (13). In particular, elevated levels of CK-BB (14) and mitochondrial CK isoenzymes (CKII) (2, 15) in the serum may cause a falsely elevated CK-MB activity even in the absence of myocardial injury (14, 16).

Studies have shown that when tumor cells enter the blood circulation and face hypoxia, CKB can be released from the cell into the extracellular space and catalyze the production of extracellular creatine phosphate, increase ATP content, maintain the energy required for tumor cell survival, and thus promote metastasis (2, 13, 17–22). Elevated circulating CKB can also interfere with CK-MB and CK activity assays, potentially explaining paradoxical CK-MB > CK patterns observed in cancer patients (23). While sporadic case reports have documented such anomalies, the biological origin and clinical relevance remain poorly characterized (24–27). Our institution, the largest cancer-specialized hospital in Southwest China, frequently observes this abnormal pattern in cancer patients. However, it remains unclear whether this is primarily driven by elevated CKB or other isoforms (28, 29).

Currently, the regulatory mechanisms underlying hematogenous metastasis of tumor cells remain poorly understood. This process is highly inefficient—fewer than 0.1% of tumor cells that enter the bloodstream ultimately succeed in forming metastatic colonies, largely due to environmental shifts and stressors such as hypoxia (17, 30). As the bloodstream serves as the primary route for cancer cells to disseminate from the primary site to distant organs, understanding how tumor cells interact with cellular and acellular components in the circulation may offer novel strategies for identifying or intercepting metastasis (30). Recent studies underscore the importance of platelet–tumor cell interactions in this process (31–34). Circulating tumor cells (CTCs) can bind to and internalize platelet components, coopting the platelet transcriptome and proteome to gain metastatic advantage (35). Tumor educated platelets (TEPs)— platelets reprogrammed by the tumor microenvironment—may facilitate tumor metastasis by secreting or transferring bioactive molecules (36–39). As the second most abundant cell population in the bloodstream, it remains unclear whether platelets can promote tumor hematogenous metastasis through factors such as CKB.

Our previous preliminary research indicated that colorectal cancer patients who developed CK-MB > CK abnormalities after curative resection exhibited significantly poorer prognoses compared to contemporaneous patients without such anomalies, characterized by shorter overall survival and a higher risk of recurrence and metastasis (40). A deeper investigation into the factors contributing to the CK-MB > CK abnormality, and the role played by tumor educated platelets in this process, may offer new insights into the mechanisms of hematogenous tumor metastasis.

In this study, we conducted a comprehensive investigation integrating retrospective cohort analysis, serum CK isoenzyme electrophoresis, and transcriptomic profiling of paired serum and platelet samples. In addition, we leveraged public RNA sequencing datasets to validate the involvement of platelet-associated CKB in lung cancer and to explore its potential role in metastatic dissemination. Our goal was to elucidate the cellular source and clinical im-plications of CK-MB > CK abnormalities, with a particular focus on the interplay between CK isoenzyme dysregulation, platelet biology, and the metastatic phenotype in cancer.

## 2. Materials and methods

### 2.1. Patient enrollment and data collection

This study combined retrospective data analysis with prospective sample collection, both of which were approved by the Institutional Ethics Committee of Sichuan Cancer Hospital and exempted from informed consent (IRB No. SCCHEC-02-2022-017).

For the retrospective component, patients diagnosed with malignant tumors and hospitalized at our institution between January 1, 2019, and December 31, 2022, were reviewed. Inclusion criteria required that patients had undergone serum CK-MB and total CK activity testing and exhibited CK-MB > CK abnormalities. The following exclusion criteria were applied: (1) presence of severe skeletal muscle injury or major cardiovascular diseases (e.g., acute myocardial infarction, myocarditis); (2) serum samples showing hemolysis or severe lipemia; (3) age younger than 18 or older than 100 years; (4) incomplete clinical data. Clinical information including sex, age, diagnosis, TNM stage, laboratory indices (CK-MB and total CK activity, platelet-related test results), treatment modalities (chemotherapy, radiotherapy, immunotherapy, targeted therapy), metastasis status, and survival outcomes was extracted from the electronic medical record system. Overall survival (OS) was defined as the time from the date of diagnosis to the date of death or the last follow-up. Retrospective data were accessed for research purposes between January 1, 2023, and May 31, 2023. All data were anonymized prior to analysis, and no personally identifiable information was accessible to the researchers.

For the prospective component, fresh serum and platelet samples were collected from a subset of colorectal and lung cancer patients admitted between April 1, 2021, and May 31, 2022. Samples were residual clinical specimens obtained during routine diagnostic testing and were anonymized prior to research use.

This study was conducted in accordance with the Declaration of Helsinki and complied with institutional and international ethical guidelines for human research. All data were independently reviewed and verified by two researchers.

### 2.2. Serum sample collection and laboratory testing

Peripheral venous blood samples were collected using BD Vacutainer tubes with clot activator and gel separator (BD, USA). Samples were centrifuged at 4,000 rpm for 8 minutes, and the supernatant serum was carefully transferred into RNase-free Eppendorf tubes and immediately stored at −80°C. Hemolytic, icteric, or lipemic samples were excluded from the analysis.

Serum total CK and CK-MB activities were measured using the phosphocreatine substrate method and immunoinhibition assay, respectively, on a Mindray BS-2000M automated biochemical analyzer (Mindray, China). The reference range for CK-MB was ≤25 U/L. The reference range for total CK was 24–194 U/L in males and 24–170 U/L in females. An abnormal result was defined as a CK-MB/CK ratio greater than 1.

Quantitative analysis of CK isoenzyme subtypes in serum samples was performed using the semi-automated HYDRASYS electrophoresis system and the HYDRAGEL ISO-CK kit (Sebia, France). Electrophoresis was conducted on an agarose gel in an alkaline buffer (pH 8.4). Densitometric analysis was performed after electrophoresis to obtain the relative quantification of each CK isoenzyme fraction.

In this study, we defined a novel composite index termed CK-Sub, representing the sum of non-CK-MM and non-CK-MB isoenzymes detected by serum isoenzyme electrophoresis. Specifically, CK-Sub was calculated as the combined relative abundance of CK-BB, CKII, and CKI.

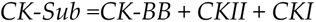

This composite measure was designed to reflect the overall contribution of non-cardiac, non-skeletal muscle CK isoenzymes, and to better capture the atypical CK isoenzyme activation patterns observed in cancer patients with CK-MB > CK abnormalities (defined as CK-MB/CK ratio abnormalities).

### 2.3. Platelet testing, separation and purity verification

First, venous blood was collected using EDTA anticoagulation tubes (BD, USA), and platelet-related parameters, including platelet count (PLT), mean platelet volume (MPV), and platelet distribution width (PDW), were measured using BC-5390 and BC-6800 fully automatic hematology analyzers (Mindray, Shenzhen). The instrument was internally quality controlled daily and calibrated weekly, and all results met the acceptable standards.

Then, whole blood samples (1.5 mL), anticoagulated with EDTA, were transferred into 2 mL microcentrifuge tubes. To obtain platelet-rich plasma (PRP), samples were centrifuged at 120 × g for 20 minutes (Shuke Instruments, Sichuan, China), effectively separating platelets from nucleated cells. To reduce contamination from leukocytes and erythrocytes, only the upper 80% of the PRP layer was carefully aspirated for subsequent procedures. Platelets were then pelleted by a second centrifugation step at 360 × g for 20 minutes. All isolation steps were completed within 2 hours of venipuncture to prevent RNA degradation. Platelet purity was assessed using morphological examination and automated platelet counts, and samples were considered acceptable if leukocyte contamination was below 5 white blood cells per 1 million platelets.

### 2.4. Quantitative reverse transcription PCR

Primer sequences were designed and synthesized by Tsingke Biological Technology Co., Ltd. (Beijing, China). The relative gene expression was quantified using the ΔCT method, with GAPDH serving as the internal control. The primer sequences used in this study were as follows: GAPDH (forward: GGAGCGAGATCCCTCCAAAAT; reverse: GGCTGTTGTCATACTTCTCATGG), CKB (forward: GCTGCGACTTCAGAAGCGA; reverse: GGCATGAGGTCGTCGATGG), CKMT1 (forward: TTGATTGTGAACGGCGTCTG; reverse: ATGCTTGGTGTGGATGACAG), and CKMT2 (forward: AGGTGACACCCAACGGCTA; reverse: TGACGGGGTCAAAAAGGTCAG).

All RT-qPCR amplification experiments were performed on a LightCycler 480 system (Roche Diagnostics). The reactions utilized the TB Green Premix Ex Taq II kit (TaKaRa, Dalian, China) according to the manufacturer’s instructions. Each assay was carried out in triplicate for biological validation. Reactions were set up in a total volume of 20 μL, consisting of 10 μL of SYBR Green Master Mix, 0.8 μL of each forward and reverse primer, 2 μL of cDNA template, and 6.4 μL of RNase-/DNase-free water. The thermal cycling conditions were: initial denaturation at 95°C for 2 minutes and 30 seconds, followed by 40 cycles of denaturation at 95°C for 5 seconds and annealing/extension at 60°C for 30 seconds. Primer specificity was confirmed by observing a single peak in the melting curve analysis.

### 2.5. Data acquisition from public repositories

Transcriptomic datasets were obtained from the Gene Expression Omnibus (GEO) database (https://www.ncbi.nlm.nih.gov/geo/). The dataset GSE89843 contains RNA-sequencing profiles of tumor-educated platelets (TEPs) from 402 non-small cell lung cancer (NSCLC) patients and 377 control individuals. The dataset GSE123902 includes single-cell RNA-seq data from clinical specimens across different disease stages: non-tumor-involved lung tissue (n = 4), primary lung adenocarcinomas (n = 8), and distant metastatic lesions located in the brain (n = 3), bone (n = 1), and adrenal gland (n = 1). The GSE74639 dataset includes RNA-Seq data of primary lung tumor cells, circulating lung tumor cells, and peripheral blood leukocytes. Additionally, bulk RNA-seq data of primary lung tumors and corresponding clinical information were retrieved from The Cancer Genome Atlas (TCGA) via the Genomic Data Commons (GDC) portal (https://portal.gdc.cancer.gov/) to assess the clinical and prognostic relevance of CKB expression in tumor tissue.

### 2.6. Statistical analysis

All statistical analyses were performed using R software (version 4.3.1; R Foundation for Statistical Computing, Vienna, Austria. http://www.r-project.org accessed on June 16, 2023). Continuous variables were presented as medians with interquartile ranges (IQRs), and categorical variables were summarized as counts and percentages. For group comparisons of continuous variables, the Wilcoxon rank-sum test was used due to non-normal data distributions.

Enrichment ratio was calculated as the proportion of each cancer type among CK-MB > CK abnormal cases divided by the proportion of that cancer type in the overall cancer patient cohort:

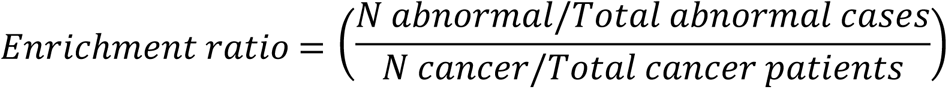

Abnormality rate for each cancer type was calculated as:

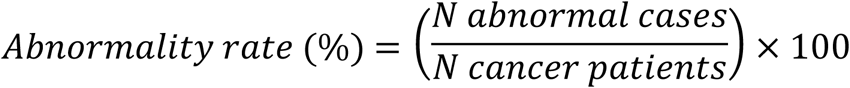

Multivariate linear regression analysis was conducted to identify independent clinical predictors of CK-MB/CK ratio, with CK-MB/CK ratio as the dependent variable, and age, sex, tumor stage, and treatment status as covariates. Treatment status was categorized as yes or no based on whether patients received any form of adjuvant therapy. Principal component analysis (PCA) was performed to explore the overall patterns of CK isoenzyme expression profiles, and PERMANOVA (permutational multivariate analysis of variance) was used to assess statistical differences between groups. Linear regression analysis was performed to assess the association between CK-Sub levels and CK-MB/CK ratio. Patients were stratified into high and low expression groups based on the optimal cutoff determined by the Youden index from receiver operating characteristic (ROC) curve analysis. Survival analyses were conducted using Kaplan-Meier curves, and differences between survival distributions were compared using the log-rank test.

To explore the potential involvement of platelets in the development of abnormal CK isoenzyme profiles, we evaluated the correlation between CK-Sub, CKBB, or CKII levels and platelet-related hematological indices, PLT, MPV, and PDW. Spearman correlation analysis was first performed to identify monotonic relationships between CK-Sub, CKBB, or CKII and each platelet parameter. Subsequently, multivariate linear regression models were constructed with CK-Sub, CKBB, or CKII as the dependent variable, adjusting for potential confounders such as tumor type, TNM stage, age, and sex. The variance inflation factor (VIF) was calculated to assess multicollinearity.

Gene expression levels were quantified as ΔCT values, and relative expression was calculated using the 2^-ΔCT method. For paired samples (i.e., matched serum and platelet samples from the same patient), the Wilcoxon signed-rank test was used to compare gene expression between compartments. If the data met normality and homogeneity assumptions, a paired t-test was applied instead. For unpaired comparisons (e.g., platelet or serum expression between colorectal and lung cancer patients), the Mann–Whitney U test or independent-samples t-test was employed depending on the distribution of the data.

Raw gene expression matrices were retrieved and processed using the GEOquery package (v2.66.0) and limma (v3.56.2). Normalized CKB mRNA expression levels were extracted and, where necessary, transformed into log₂(TPM + 1). Statistical comparisons between groups were conducted using the Wilcoxon rank-sum test or paired Wilcoxon signed-rank test, as appropriate.

TCGA-LUAD RNA sequencing data and corresponding clinical survival information were accessed via the UCSC Xena browser. Patients were stratified into high and low CKB expression groups based on the median CKB expression level. Kaplan-Meier survival curves were plotted, and log-rank tests were used to assess statistical significance. Hazard ratios (HRs) and 95% confidence intervals (CIs) were estimated using Cox proportional hazards regression analysis.

For differential gene expression (DEG) analysis of platelet RNA-seq data from lung cancer patients, the DESeq2 package (v1.36.0) was applied. Genes significantly correlated with platelet CKB expression (adjusted p < 0.05 and |log₂FC| > 1) were intersected with CKB-associated DEGs from tumor tissues. Functional enrichment analyses, including Gene Ontology (GO) and Kyoto Encyclopedia of Genes and Genomes (KEGG) pathway analysis, were performed using the clusterProfiler package (v4.4.4). Data visualization, including expression distribution and group comparisons, was performed using ggplot2 (v3.4.2).

A two-tailed P-value < 0.05 was considered statistically significant for all analyses.

## 3. Results

At first, patients with CK-MB > CK anomalies were retrospectively identified from a large-scale cancer cohort, and cancer types with high prevalence or enrichment were characterized. Then, CK isoenzyme profiles in serum were analyzed using electrophoresis and qRT-PCR to identify cancer-specific patterns. Next, correlations between platelet parameters and CK isoenzymes were explored, and the expression of CKB in paired serum and platelet samples was quantitatively validated. Finally, public transcriptomic datasets were utilized to confirm CKB overexpression in lung cancer platelets, investigate its metastatic relevance, and elucidate potential functional pathways. The overall workflow and key findings were illustrated in Fig 1.

**Fig 1.**
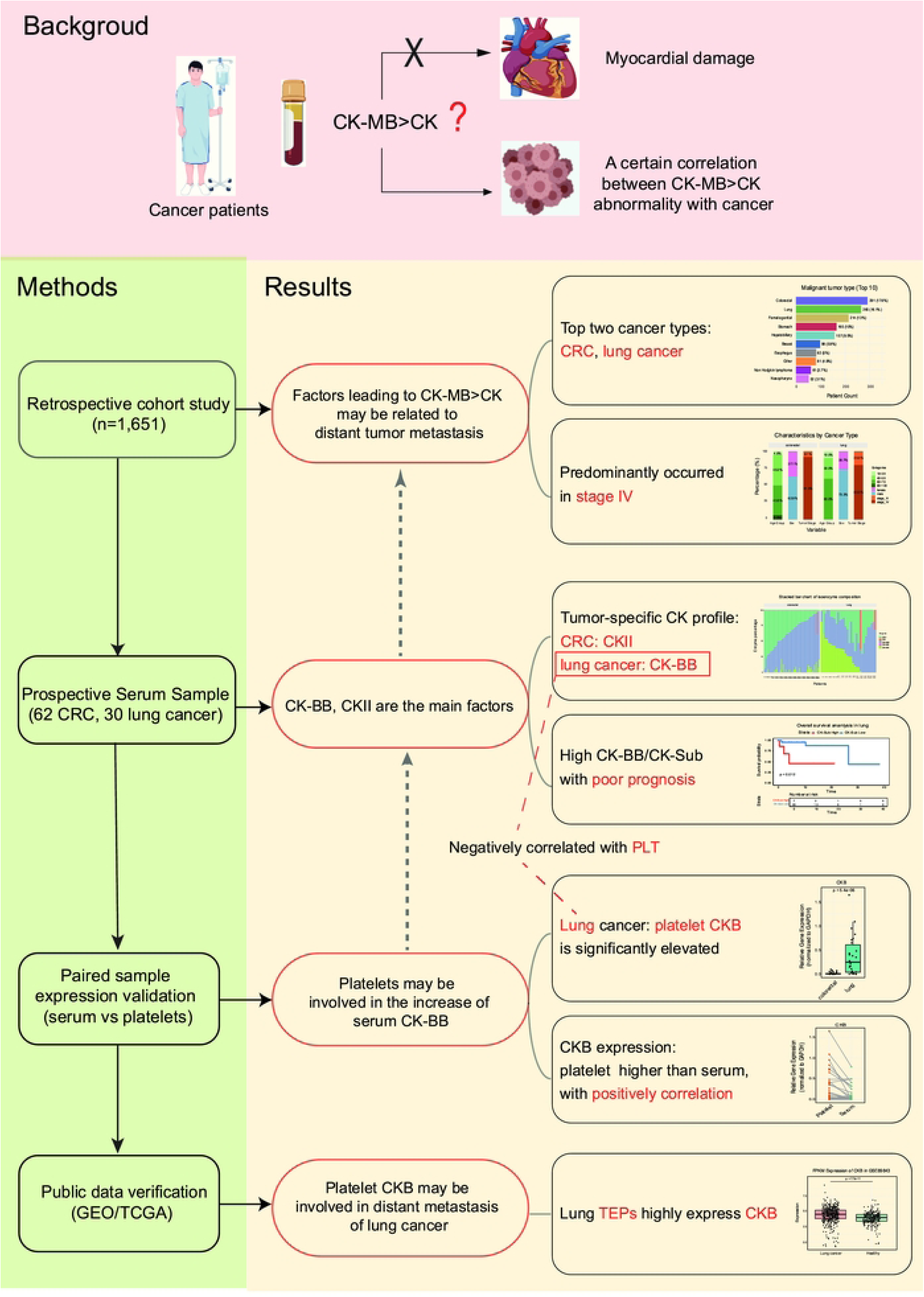
Study workflow and key findings. Background: In cancer patients, the clinical association between serum CK-MB > CK abnormality and tumors is unclear. Methods: First, a large cancer cohort was screened to characterize the epidemiological distribution of CK-MB > CK abnormality. Then, isozyme analysis and qRT-PCR were used to identify cancer type-specific expression of CK isozymes in serum and platelets. Further, the expression of platelet CK was validated using a public TEP dataset (GSE89843), and single-cell RNA sequencing data (GSE123902) were analyzed to explore the role of CKB in lung cancer metastasis. Results: CKB was identified as a platelet-derived transcript enriched in lung cancer patients with elevated serum CK-BB levels. Platelet CKB expression was associated with distant metastasis and poor prognosis, suggesting its potential functional role in shaping the metastatic tumor microenvironment. Abbreviations: CK-MB, creatine kinase-MB isoenzyme; CK, creatine kinase; CRC, colorectal cancer; CK-BB, creatine kinase brain-type isoenzyme; CKII, mitochondrial creatine kinase isoenzyme II; PLT, platelet count; TEPs, tumor-enhanced platelets.

### 3.1 Patient characteristics and clinical correlates of CK-MB > CK abnormalities

A total of 1,651 cancer patients presenting with CK-MB > CK abnormalities were retrospectively enrolled between 2019 and 2022. Most patients were aged between 40 and 79 years (Fig 2A). Based on ICD codes (S1 Table), colorectal cancer (291 cases, 17.6%), lung cancer (265 cases, 16.1%), and female reproductive system tumors (214 cases, 13.0%) were the top three cancer types represented (Fig 2B). To correct for differences in overall hospitalization frequencies across cancer types, we normalized CK-MB > CK occurrence against the total number of patients per cancer type. This revealed that colorectal cancer exhibited the highest enrichment and abnormality rate, indicating a strong association with CK-MB > CK anomalies. In contrast, the high absolute case count in lung cancer likely reflected its high population prevalence rather than specific enrichment (Fig 2C).

**Fig 2.**
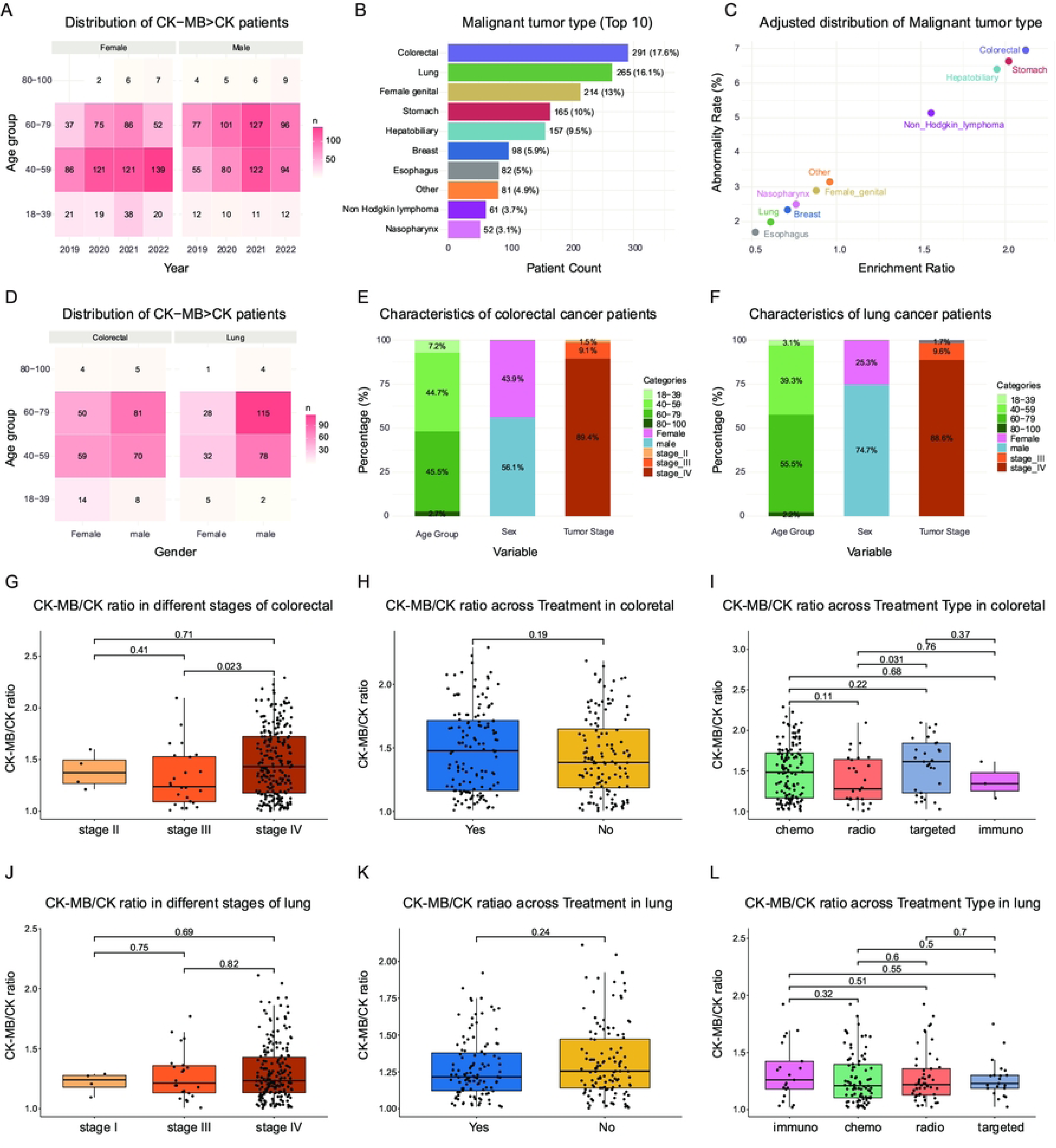
Patient characteristics and clinical correlates of CK-MB > CK abnormalities. (**A**) Age and sex distribution of patients with CK-MB > CK abnormality from 2019 to 2022. (**B**) Top 10 malignant tumor types among patients with CK-MB > CK abnormality, based on patient counts. (**C**) Adjusted distribution of tumor types after normalizing by total visits during the same period. Statistical methods: (**D**) Age and sex distribution of colorectal and lung cancer patients with CK-MB > CK abnormality. (**E**) Clinical characteristics (age group, sex, and tumor stage) of colorectal cancer patients. (**F**) Clinical characteristics (age group, sex, and tumor stage) of lung cancer patients. (**G–I**) CK-MB/CK ratio comparisons across tumor stages, adjuvant therapy status, and different treatment types in colorectal cancer patients. (**J–L**) CK-MB/CK ratio comparisons across tumor stages, adjuvant therapy status, and different treatment types in lung cancer patients. Enrichment ratio was calculated as the proportion of each cancer type among CK-MB > CK abnormal cases divided by the proportion of that cancer type in the overall cancer patient cohort. Abnormality rate was calculated as the proportion of CK-MB > CK abnormal cases within each cancer type divided by the total number of patients diagnosed with that cancer type. Statistical methods: Pairwise comparisons of CK-MB/CK ratio between groups were conducted using the Wilcoxon rank-sum test. Abbreviations: CK-MB, creatine kinase-MB isoenzyme; CK, creatine kinase; chemo, chemotherapy; radio, radiotherapy; targeted, targeted therapy; immuno, immunotherapy.

To further explore the clinical relevance of these abnormalities, we focused on colorectal and lung cancer patients. In colorectal cancer (n = 291), the majority of patients with CK-MB > CK anomalies were aged between 40–80 years (Fig 2D) and stage IV (89.4%) (Fig 2E). A significantly higher CK-MB/CK ratio was observed in stage IV compared to stage III patients (P = 0.023, Fig 2G), suggesting a potential link with tumor progression. However, no significant associations were found between CK-MB/CK ratio and treatment modality or adjuvant therapy status (Figs 2H–1I). Multivariate regression confirmed that age, sex, stage, and therapy were not independently associated with CK-MB/CK ratio levels (S2 Table).

Similarly, in the lung cancer cohort (n = 265), patients also tended to be older (Fig 2D), with a male predominance (74.7%) and high proportion of advanced-stage disease (88.6% stage IV, Fig 2F). No significant associations were observed between CK-MB/CK ratio and tumor stage, adjuvant therapy, or treatment modality (Figs 2J–1L), and multivariate analysis again showed no independent clinical predictors of CK-MB/CK ratio elevation (S2 Table), suggesting that additional factors may be involved in explaining the observed CK-MB > CK abnormalities.

### 3.2. Creatine kinase isoenzyme profile analysis in colorectal and lung cancer patients

Isoenzyme electrophoresis was performed prospectively on 92 serum samples (62 colorectal, 30 lung cancer patients) exhibiting CK-MB > CK abnormalities (Fig 3A). All patients were stage III/IV (Fig 3B), consistent with the retrospective cohort, further suggesting that CK-MB > CK abnormalities were closely associated with enhanced tumor aggressiveness. Electrophoretic analysis revealed consistent abnormal bands across samples, predominantly involving elevated CK-BB and CKII isoenzymes, with most patients displaying a bimodal peak pattern (Fig 3C). Further comparison between colorectal and lung cancer patients showed that CKII elevation predominated in colorectal cancer, whereas both CKII and CK-BB elevations were observed in lung cancer (Fig 3D).

**Fig 3.**
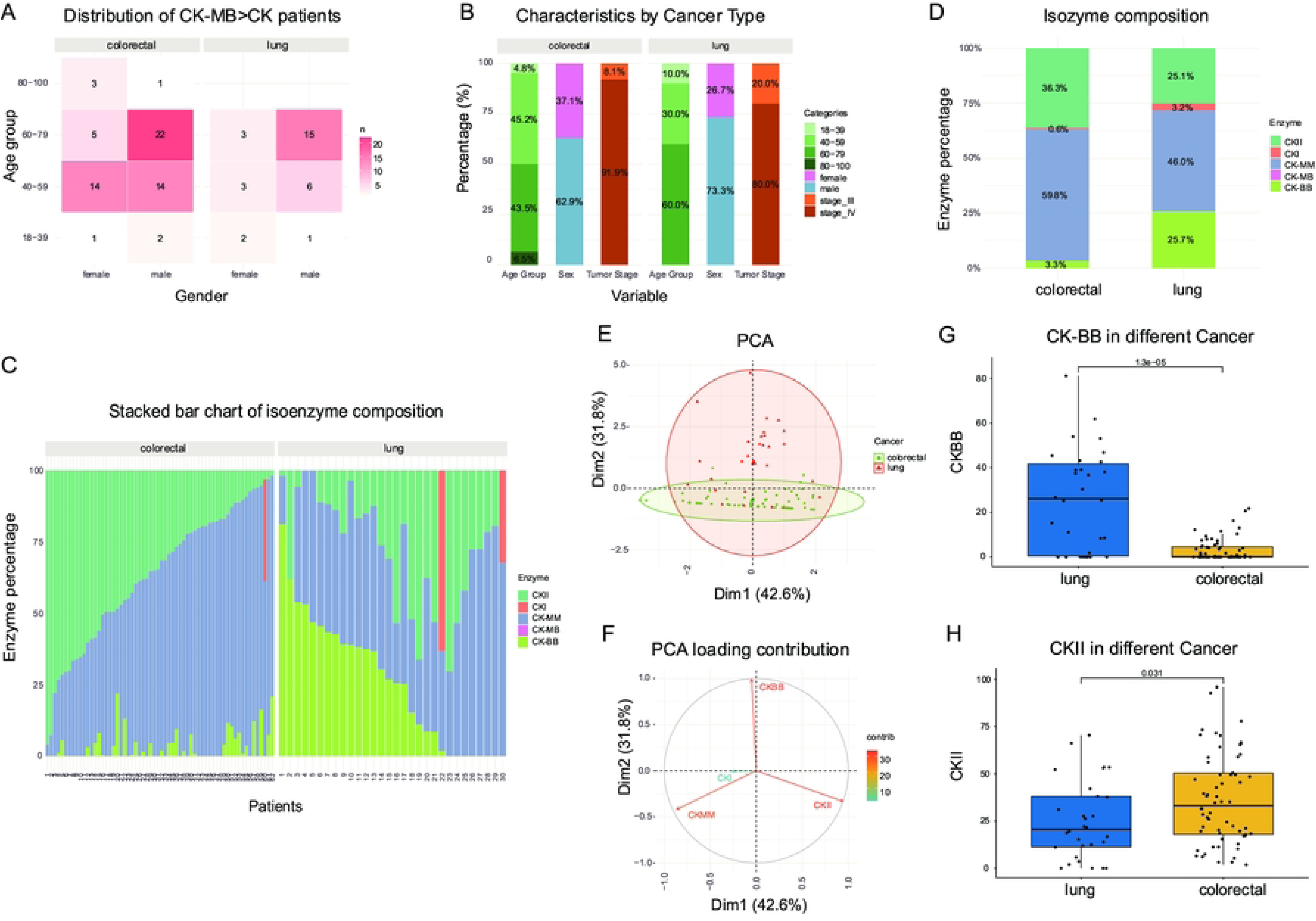
Isoenzyme profile analysis in colorectal and lung cancer patients with CK-MB > CK abnormality. (**A**) Age and sex distribution of colorectal and lung cancer patients included in the isoenzyme electrophoresis analysis. (**B**) Clinical characteristics of colorectal and lung cancer patients. (**C**) Stacked bar plot of CK isozyme composition for each patient. (**D**) Overall isoenzyme composition by cancer type. (**E**) Principal component analysis (PCA) showing separation of colorectal and lung cancer patients based on isoenzyme expression profiles. (**F**) PCA loading plot indicating the contribution of different CK isoenzymes (CK-BB, CKII, CK-MB, CK-MM, CKI) to the variance. (**G-H**) Comparison of CK-BB and CKII expression between lung cancer and colorectal cancer patients. Statistical methods: Pairwise comparisons were conducted using the Wilcoxon rank-sum test. Abbreviations: CK, creatine kinase; CK-MB, creatine kinase-MB isoenzyme; CK-BB, creatine kinase brain-type isoenzyme; CK-MM, creatine kinase muscle-type isoenzyme; CKII, mitochondrial creatine kinase isoenzyme II; CKI, mitochondrial creatine kinase isoenzyme I; PCA, principal component analysis.

PCA showed that colorectal cancer samples clustered along Dim1, associated with increased CKII expression, whereas lung cancer samples clustered along Dim2, associated with CK-BB elevation (PERMANOVA, P=0.001) (Figs 3E–2F). Comparative analysis confirmed that lung cancer patients had significantly higher CK-BB levels (P=1.3e-5) and lower CKII levels (P=0.031) compared to colorectal cancer patients (Figs 3G–2H), suggesting cancer type-specific CK isoenzyme heterogeneity.

### 3.3. Clinical relevance of CK isoenzyme alterations in colorectal and lung cancer

To investigate the clinical implications of serum CK isoenzyme abnormalities in cancer patients with CK-MB > CK patterns, we conducted a stratified analysis of colorectal cancer (n=62) and lung cancer (n=30) patients who underwent isoenzyme electrophoresis.

In colorectal cancer patients, linear regression analysis revealed a mild positive linear relationship between CK-Sub (CK-Sub =CK-BB + CKII + CKI) levels and CK-MB/CK ratio (R² = 0.098), although the explanatory power was limited, suggesting that variations in CK-Sub alone may be insufficient to effectively predict CK-MB/CK ratio abnormalities (Fig 4A). We found that stage IV patients exhibited significantly higher CKII (P = 0.023) and CK-Sub (P = 0.022) levels compared with stage III patients (Figs 4C–3D), indicating that specific isoenzyme upregulation may be linked to tumor progression. In contrast, no significant differences were found in CK-BB expression (P = 0.41) or CK-MB/CK ratio (P = 0.86) between stages (S1 Fig), suggesting that the CK-MB/CK ratio alone may not sensitively reflect tumor advancement. Kaplan-Meier survival analysis revealed that patients with high CKII expression had significantly shorter overall survival (Fig 4F). Similarly, elevated CK-Sub levels were associated with worse survival outcomes (HR, 0.17; 95%CI, 0.0421-0.688; P = 0.0049) (Fig 4G). Although the overall variation in CK-MB/CK ratio was limited, alterations in individual isoenzyme components, particularly CKII, or the total CK-Sub, may serve as potential biomarkers for tumor progression in advanced-stage colorectal cancer.

**Fig 4.**
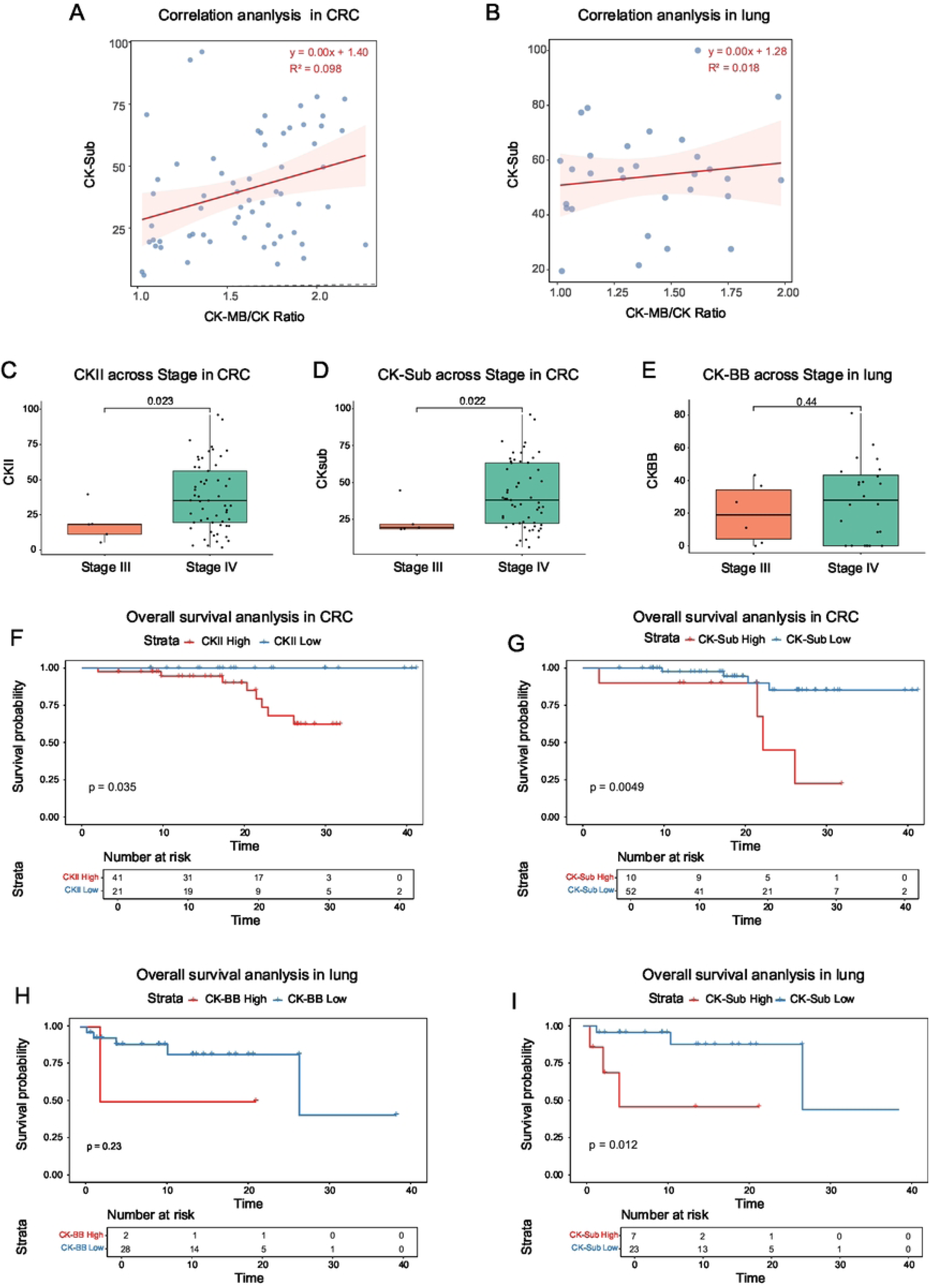
Serum CK isoenzyme alterations and clinical relevance in colorectal and lung cancer patients with CK-MB > CK abnormality. (**A**) Linear regression analysis between CK-Sub levels and CK-MB/CK ratio in colorectal cancer patients. (**B**) Linear regression analysis between CK-Sub levels and CK-MB/CK ratio in lung cancer patients. (**C-D**) Comparison of CKII, and CK-Sub between stage III and stage IV colorectal cancer patients. (**E**) Comparison of CK-BB expression between stage III and stage IV lung cancer patients. (**F–G**) Kaplan-Meier survival analyses of overall survival according to CKII and CK-Sub levels in colorectal cancer patients. (**H–I**) Kaplan-Meier survival analyses of overall survival according to CK-BB and CK-Sub expression levels in lung cancer patients. Statistical methods: Linear regression analysis was used to assess the association between CK-MB/CK ratio and CK-Sub levels. R² value represents the proportion of variance in CK-Sub explained by CK-MB/CK ratio. Group comparisons were performed using the Wilcoxon rank-sum test. Overall survival was analyzed by Kaplan-Meier curves, and differences between groups were assessed with the log-rank test. Abbreviations: CK-MB, creatine kinase-MB isoenzyme; CK, creatine kinase; CKII, mitochondrial creatine kinase isoenzyme II; CK-BB, creatine kinase brain-type isoenzyme; CK-Sub, total CK isoenzyme activity (CK-BB + CKII + CKI); CRC, colorectal.

Unlike the mild positive linear trend observed in colorectal cancer patients, linear regression analysis in lung cancer patients revealed no significant linear relationship between CK-Sub levels and CK-MB/CK ratio (Fig 4B). In the comparison across tumor stages, CK-BB levels were higher in stage IV patients; however, the difference did not reach statistical significance (P = 0.44) (Fig 4E), possibly due to the limited sample size. Similarly, no significant differences were observed in CKII, CK-Sub levels, or CK-MB/CK ratio between different tumor stages (S1 Fig). Regarding survival outcomes, although CK-BB levels did not significantly predict overall survival (P=0.23) (Fig 4H), a general trend toward poorer survival in the high-expression group was noted. Notably, patients with high CK-Sub levels had significantly shorter overall survival (P = 0.012) (Fig 4I). These findings suggest that tumor progression in lung cancer may involve abnormal activation of multiple CK isoenzyme subtypes. Although individual isoenzyme markers may not independently predict prognosis due to sample size limitations, the overall increase in total CK isoenzyme activity (CK-Sub) appears to be closely associated with poorer clinical outcomes.

### 3.4 Association between platelet parameters and CK isoenzymes

To investigate whether platelets are involved in the aberrant expression patterns of CK isoenzymes observed in cancer patients, we first assessed the correlations between routine platelet-related hematological parameters (including PLT, MPV, and PDW) and serum CK isoenzyme indices (CK-Sub, CK-BB, and CKII) in patients with colorectal and lung cancers. After adjusting for potential confounding variables such as cancer type, tumor stage, age, and sex using multiple linear regression, we found a significant inverse association between PLT and serum CK-BB levels (β = –0.038, p = 0.034). Variance inflation factor (VIF) analysis confirmed the absence of multicollinearity among covariates (all VIFs < 2.5), supporting the robustness of the model (Table 1). These findings suggest that reduced PLT may be closely linked to elevated serum CK-BB levels. Notably, upon inclusion of an interaction term (PLT × Cancer) in the regression model, the interaction between PLT and lung cancer approached statistical significance (β = –0.055, p = 0.053), indicating a potentially stronger inverse association between PLT and CK-BB expression specifically in lung cancer patients.

**Table 1.**
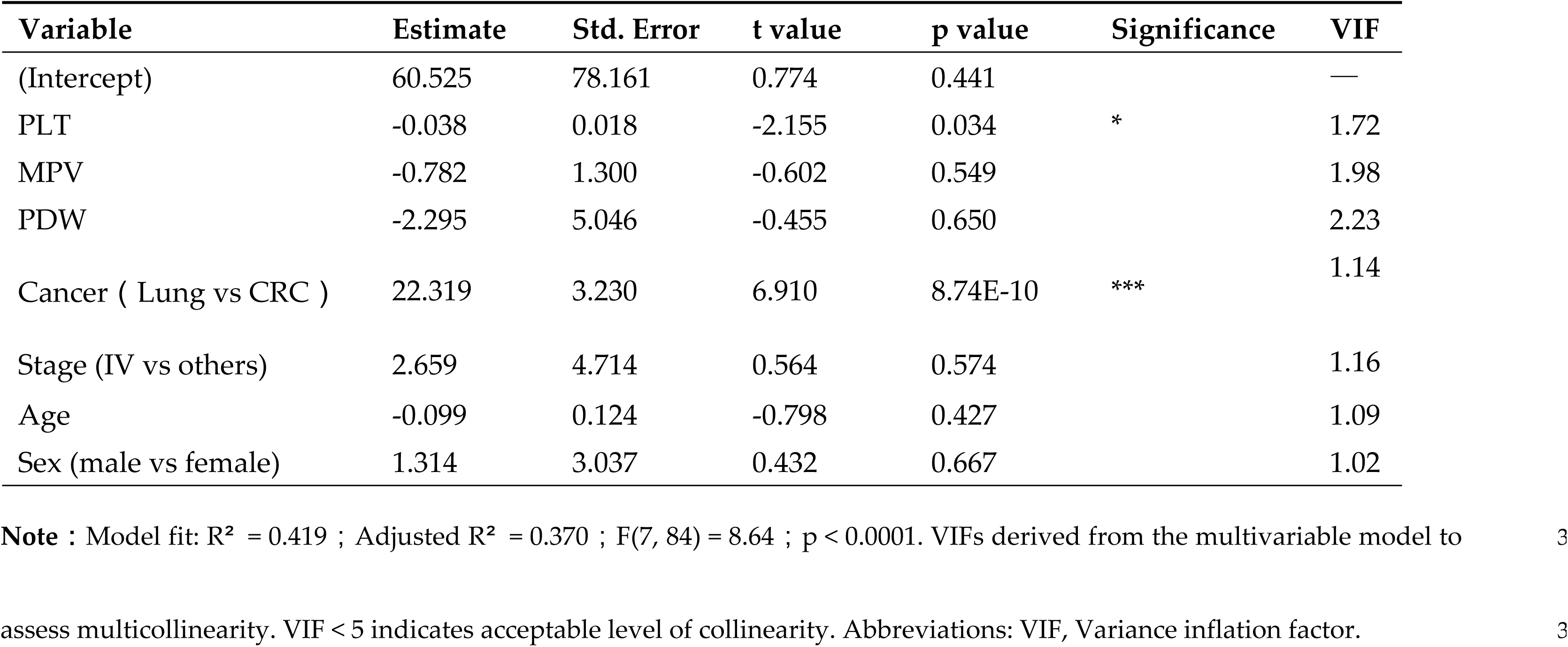
Multiple Linear Regression Analysis with VIF of Serum CK-BB Expression and Platelet Parameters.

3.5 ***Quantitative RT-PCR detection of CK family genes in serum and platelets***

Furthermore, we performed qRT-PCR analysis to quantify the mRNA expression levels of three creatine kinase genes—CKB, CKMT1, and CKMT2—in paired serum and platelet samples collected from cancer patients. The expression profiles in serum closely mirrored the protein isoenzyme patterns observed in previous electrophoretic analyses, exhibiting marked cancer-type specificity: mitochondrial isoenzyme CKMT1 was predominantly expressed in colorectal cancer, whereas CKB expression was markedly elevated in lung cancer (Figs 5A–4B). All three CK isoenzyme transcripts were detectable in the paired platelet samples, indicating active transcription of CK genes in platelets. Notably, platelet-derived CKB mRNA expression was significantly higher in lung cancer patients compared to those with colorectal cancer (p = 5.4 × 10⁻⁶; Fig 5C), suggesting potential cancer-type-specific upregulation in the lung cancer context. Paired sample analysis further revealed that, in the majority of cases, platelet CKB mRNA levels exceeded the corresponding serum expression levels (p = 0.03, Wilcoxon signed-rank test; Fig 5D). Additionally, correlation analysis demonstrated a significant positive association between serum and platelet CKB expression levels (R² = 0.33, p = 2.9 × 10⁻⁵; Fig 5E). Collectively, these findings suggest that platelets may serve as a critical source of serum CKB in cancer patients—particularly in lung cancer—potentially contributing to the observed CK-MB > CK isoenzyme elevation phenotype.

**Fig 5.**
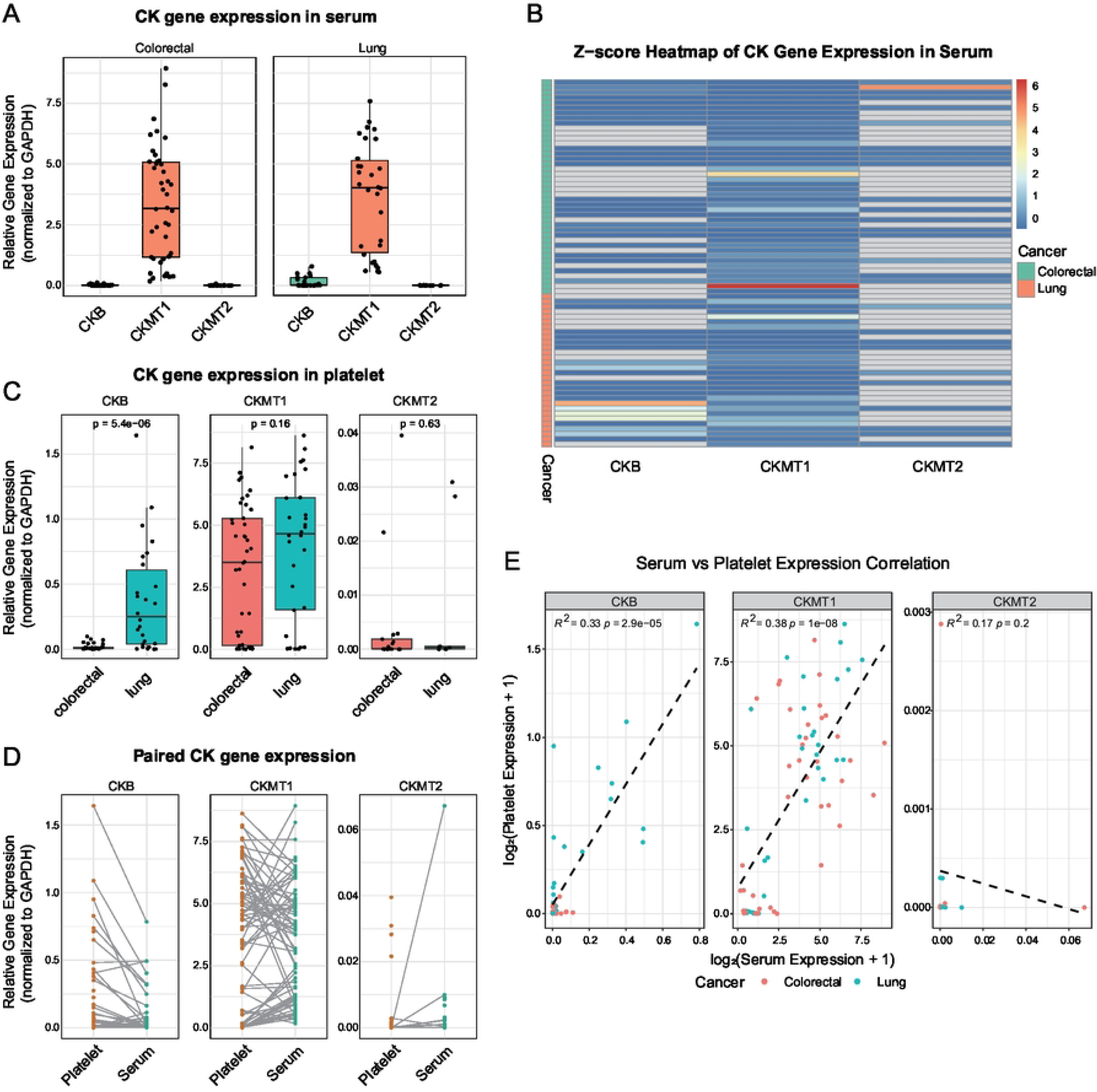
Differential expression and correlation of CK genes in serum and platelets. **(A)** Expression of CKB, CKMT1, and CKMT2 mRNA in serum samples from lung and colorectal cancer patients. **(B)** Z-score heatmap displaying CK genes expression patterns in serum stratified by cancer type. **(C)** Comparison of platelet CK genes expression between colorectal and lung cancer patients. **(D)** Paired expression analysis of CK genes in platelet versus serum from the same patients. **(E)** Correlation analysis between serum and platelet CK genes expression. Statistical methods: Wilcoxon rank-sum test was used for comparisons between colorectal and lung cancer groups in panels. Paired Wilcoxon signed-rank test was used for within-patient comparisons between serum and platelet expression. Pearson correlation analysis was used to evaluate association between serum and platelet mRNA expression levels. Expression values were log₂-transformed prior to analysis. Abbreviations: CK, creatine kinase.

### 3.6 Potential role of platelet CKB in distant metastasis of lung cancer

To further validate whether CKB is upregulated in the platelets of lung cancer patients, we analyzed RNA sequencing data from the tumor-educated platelet (TEP) dataset GSE89843. The results showed that CKB mRNA expression was significantly elevated in the platelets of lung cancer patients compared to healthy individuals (P = 7.7 × 10⁻¹¹, Fig 6A). Using the single-cell RNA sequencing dataset GSE123902, we assessed the expression of CKB mRNA within the tumor microenvironment. Notably, tumor cells from distant metastatic lesions in stage IV lung cancer patients exhibited markedly higher CKB expression, whereas no elevation was observed in primary tumor lesions (Figs 6D–5G). This finding is consistent with TCGA data, which demonstrated that while CKB expression is generally low in primary lung tumor tissues, patients with relatively higher tumor CKB expression had significantly worse survival outcomes (Figs 6C, 5H). These results collectively suggest that elevated CKB expression may serve as a potential promoter of distant metastasis.

**Fig 6.**
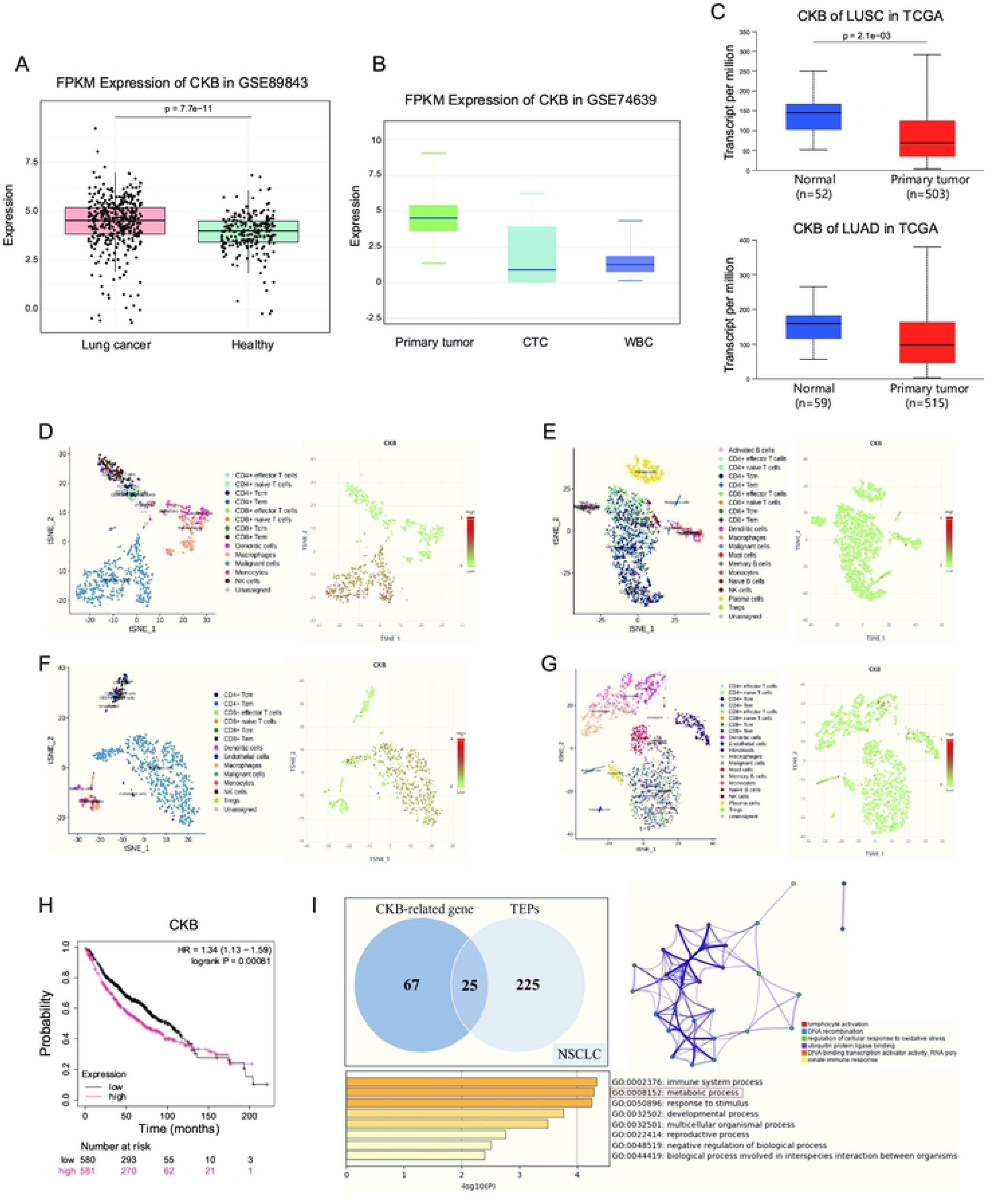
Platelet CKB expression and its potential association with lung cancer metastasis. (**A**) CKB mRNA is significantly upregulated in platelets from lung cancer patients (GSE89843); (**B**) CKB mRNA expression is lower in circulating tumor cells (CTCs) and peripheral blood leukocytes (WBCs) compared to primary tumor cells (GSE74639); (**C**) CKB mRNA is expressed at low levels in primary lung tumor tissues (TCGA dataset); (**D, F**) Tumor cells from distant metastatic lesions in stage IV lung cancer patients exhibit high CKB mRNA expression (GSE123902); (**E, G**) Tumor cells from primary lesions in stage IV lung cancer exhibit low CKB mRNA expression (GSE123904); (**H**) Lung cancer patients with high tumor CKB expression show poorer overall survival (TCGA data); (**I**) Functional enrichment and interaction analysis of CKB-associated genes in platelets. Statistical methods: Expression comparisons were performed using Wilcoxon rank-sum tests; Survival analysis was conducted using Kaplan–Meier curves and log-rank tests; Differentially expressed genes were identified using |log₂FC| > 1 and FDR < 0.05 as thresholds; Functional enrichment was assessed using clusterProfiler with GO Biological Process and KEGG pathway databases. Abbreviations: CKB, brain-type creatine kinase; NSCLC, non-small cell lung cancer; LUSC, Lung Squamous Cell Carcinoma; LUAD, Lung Adenocarcinoma; TEP, tumor-educated platelet; CTC, circulating tumor cell; WBC, White Blood Cell; GO, Gene Ontology; TCGA, The Cancer Genome Atlas.

**Fig 7.**
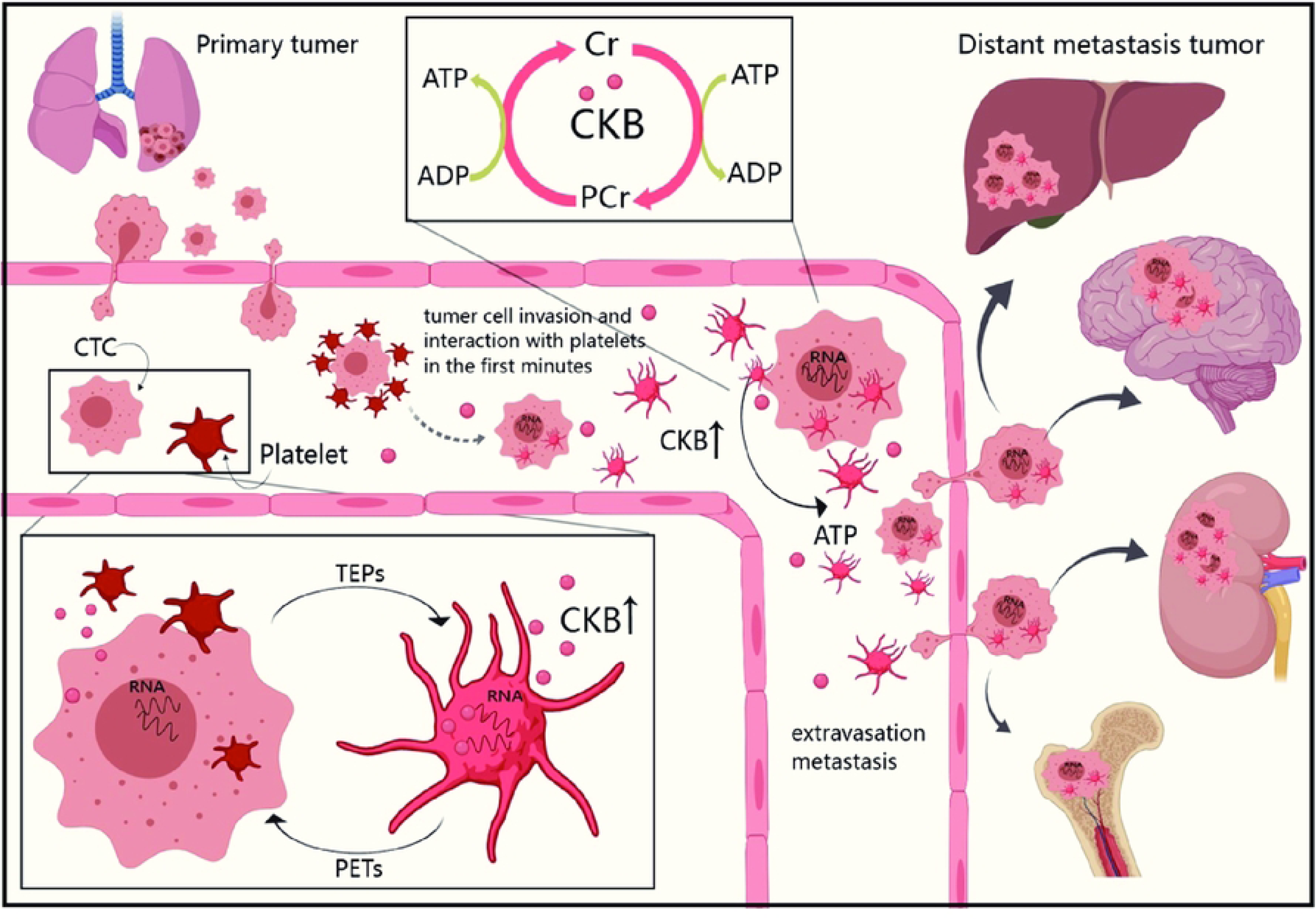
Proposed mechanism of platelet CKB in promoting hematogenous metastasis of lung cancer. Upon vascular invasion, circulating tumor cells (CTCs) rapidly interact with platelets. In response to tumor-derived signals, platelets become tumor-educated platelets (TEPs), characterized by increased CKB mRNA expression. Through intercellular exchange, tumor cells may acquire of exogenous CKB, which facilitates the generation of phosphocreatine (PCr) from creatine (Cr), maintaining ATP supply under hypoxic or shear stress conditions. This metabolic support may enhance CTC survival in circulation and promote distant organ colonization. CKB expression is elevated in metastatic tumor lesions but not in primary tumors, suggesting a potential role in metastasis initiation. PETs: platelet-educated tumor cells.

To explore the origin of elevated circulating CKB, we used GSE74639 data to analyze CKB mRNA levels in circulating tumor cells (CTCs) and peripheral blood leukocytes. Both cell populations showed markedly lower CKB expression than primary tumors (Fig 6B). Taken together with the high expression of CKB mRNA observed in peripheral platelets from stage IV patients, we speculated that platelets may represent an external source driving CKB accumulation in metastatic lung cancer cells.

To further explore the potential mechanisms by which platelet-derived CKB contributes to lung cancer metastasis, we performed an integrative analysis of CKB-associated differentially expressed genes (DEGs) in lung tumor tissues and platelet RNA profiles. A total of 25 overlapping DEGs were identified. Gene Ontology (GO) and KEGG pathway enrichment analysis revealed that these genes were significantly enriched in pathways related to metabolic processes, oxidative stress responses, immune regulation, and intercellular signaling (Fig 6I). These findings provide preliminary mechanistic insights, suggesting that platelet-derived CKB may play a functional role in shaping the metastatic tumor microenvironment and promoting distant dissemination in lung cancer.

## 4. Discussion

In this study, we comprehensively analyzed the clinical, biochemical, and transcriptomic landscape of cancer patients exhibiting the paradoxical CK-MB > CK abnormality. By integrating large-scale retrospective data, serum isoenzyme profiling, qRT-PCR analysis of platelet and serum mRNA, and bioinformatics, we uncovered that elevated CKB mRNA levels in platelets correlated with metastatic features, suggesting a possible involvement of platelet-associated CKB in lung cancer progression.

Our results showed that colorectal and lung cancers as the most frequently associated malignancies with CK-MB > CK abnormalities, particularly at advanced stages. Contrary to classical myocardial injury interpretations, electrophoresis and mRNA profiling revealed that these anomalies were primarily driven by aberrant activation of non-cardiac CK isoenzymes - namely, brain-type CK (CK-BB) and mitochondrial CK isoenzymes (CKII and CKI). To better capture these atypical patterns, we developed a composite index termed CK-Sub, defined as the sum of CK-BB, CKII, and CKI activity. This index demonstrated prognostic relevance, particularly in colorectal cancer, where elevated CK-Sub levels were significantly associated with stage IV disease and shorter overall survival. Importantly, cancer-type– specific CK expression patterns emerged: CKII was the predominant isoenzyme in CRC, while CK-BB was selectively enriched in lung cancer patients(41). Previous studies have indicated that CK-BB promotes extracellular ATP production under hypoxic conditions, thereby supporting tumor cell survival, migration, and metastasis(17, 42, 43). Our findings are consistent with these biological roles, confirming the presence of high CK-BB levels in patients with advanced-stage lung cancer(44). Notably, our study observed the important contribution of CKII to CK-MB > CK discrepancies, especially in colorectal cancer(3, 45). This highlights the potential role of mitochondrial metabolic reprogramming in tumor progression(42, 46–54)

The extracellular presence of CK-BB and mitochondrial CK may not only serve as passive markers of tumor burden but may also actively contribute to establishing a metabolically supportive microenvironment that promotes tumor survival and metastasis(55, 56). Notably, although the CK-MB/CK ratio itself exhibited limited sensitivity in predicting tumor stage or prognosis, the underlying isoenzyme signatures - particularly the elevations in CKII - showed significant correlations with disease stage. By integrating signals from multiple non-traditional CK isoenzymes, CK-Sub demonstrated significant prognostic value in both colorectal and lung cancer patients, with elevated levels indicating worse survival outcomes. These findings suggest that analyzing CK isoenzyme patterns can provide deeper biological and clinical insights compared to traditional CK-MB/CK ratio analysis(13, 19, 40), and that the cumulative activity of CK isoenzymes beyond CK-MB and CK-MM may more effectively reflect tumor aggressiveness(52).

A key and novel insight from our study is the identification of platelets as a non-canonical source of circulating CKB. qRT-PCR analysis revealed that CKB mRNA expression was significantly higher in the platelets of lung cancer patients than in colorectal cancer patients. Paired sample analysis confirmed that platelet CKB levels exceeded those in serum, and a significant positive correlation was observed between the two, indicating that platelets may actively contribute to the pool of circulating CK-BB. This hypothesis was further substantiated by analysis of the GSE89843 TEPs RNA-seq dataset, which confirmed that CKB was significantly upregulated in platelets of lung cancer patients. Additional exploration using the GSE123902 single-cell RNA-seq dataset revealed that CKB expression was specifically enriched in tumor cells within distant metastatic lesions, but not in primary tumors. These observations, combined with TCGA survival analysis linking high tumor CKB expression to poor prognosis. Functional enrichment analysis revealed that 25 shared differentially expressed genes associated with TEPs and CKB tumor related genes in lung cancer were enriched in oxidative stress response, immune regulation, and metabolic reprogramming. All evidences suggest that CKB may serve as a functional facilitator of lung cancer metastasis.

Taken together, our findings highlight a previously underexplored role of platelets as potential metabolic enablers of tumor dissemination. The discovery of elevated CKB mRNA levels in TEPs, particularly in lung cancer patients, and their correlation with distant metastasis raises the possibility that platelets may help shape the metastatic phenotype through metabolic crosstalk. We propose a mechanistic model (Fig 6) in which platelet-associated CKB supports the survival and adaptability of CTCs during hematogenous transit, potentially by enhancing ATP production in energy-stressed environments. Although our study does not provide direct evidence for the transfer or translation of platelet CKB in vivo, the convergence of clinical, biochemical, and transcriptomic findings provides a compelling rationale for further investigation.

This study also proposed that platelets may represent an important source of abnormally elevated serum CKB in cancer patients, especially in lung cancer. This not only complements the mechanistic basis for the clinical findings (CK-MB > CK), but also offers a theoretical framework for future studies on how platelets contribute to the release, transport, and systemic dissemination of tumor-associated metabolic factors such as CKB. Future studies incorporating co-culture systems, metabolic tracing, and in vivo metastasis assays are warranted to elucidate the precise functional contributions of platelet CKB in lung cancer progression. This may provide new clues for blocking platelet-assisted hematogenous metastasis.

## 5. Conclusions

This study suggests that in lung cancer patients with serum CK-MB>CK abnormalities, the atypical CK pattern is driven by CK-BB specificity. Among them, platelet CKB is significantly elevated and closely related to distant metastasis and poor prognosis. Platelet CKB may be a potential participant in hematogenous dissemination of lung cancer, and its mechanism deserves further investigation.

## Data Availability

All relevant data are within the manuscript and its Supporting Information files.

## Abbreviations

ATP: Adenosine Triphosphate
CK: Creatine Kinase
CK-MB: Creatine Kinase-MB Isoenzymes
CK-BB: Creatine Kinase Brain-Type Isoenzymes
CK-MM: Creatine Kinase Muscle-Type Isoenzymes
MtCK: Mitochondrial Creatine Kinase
CKII: Mitochondrial Creatine Kinase Isoenzyme II (dimer form)
CKI: Mitochondrial Creatine Kinase Isoenzyme I (monomer form)
RC: Colorectal cancer
PCA: Principal Component Analysis
PERMANOVA: Permutational Multivariate Analysis of Variance
HR: Hazard Ratio
CI: Confidence Interval
OS: Overall Survival
CTC: Circulating tumor cell
TEP: Tumor-educated platelet
PET: Platelet-educated tumor cell
PLT: Platelet count
MPV: Mean platelet volume
PDW: Platelet distribution width
PRP: Pplatelet-rich plasma
NSCLC: Non-small cell lung cancer
ROC: Receiver operating characteristic
VIF: Variance inflation factor
DEG: Differential gene expression
WBC: White blood cell

## Acknowledgments

During the preparation of this work, the authors used ChatGPT (OpenAI, GPT-4 version) for the purpose of assisting in refining the English language expressions. All scientific content, analyses, interpretations, and conclusions were independently developed by the authors. After using this tool, the authors have carefully reviewed and edited all outputs and take full responsibility for the content and interpretation presented in this publication.

## Supporting information

**S1 Table.** Cancer Group Classification Based on ICD-10 Codes.

| Cancer group | ICD - 10 codes | Description |
| --- | --- | --- |
| Colorectal | C18, C19, C20 | Colon, rectosigmoid junction, rectum |
| Head and neck | C00–C06, C07–C08, C09–C10, C12–C14 | Lip, oral cavity, salivary glands, pharynx |
| Hepatobiliary | C22, C23, C24 | Liver and biliary system |
| Female genital | C51–C57 | Vulva, vagina, cervix, corpus uteri, ovary, etc. |
| Urinary system | C64–C68 | Kidney, renal pelvis, ureter, bladder, etc. |
| Secondary_metastatic | C77, C78, C79 | Lymph node, respiratory and digestive metastasis |
| Non Hodgkin lymphoma | C82, C83, C84, C85 | Different types of non-Hodgkin lymphoma |
| Hematologic malignancy | C88, C90–C96 | Blood malignancies (multiple myeloma, leukemia, etc.) |
| Lung | C34 | Lung cancer |
| Stomach | C16 | Gastric cancer |
| Esophagus | C15 | Esophageal cancer |
| Breast | C50 | Breast cancer |
| Prostate | C61 | Prostate cancer |
| Nasopharynx | C11 | Nasopharyngeal cancer |
| Pancreas | C25 | Pancreatic cancer |
| Brain | C71 | Brain tumors |
| Thyroid | C73 | Thyroid carcinoma |
| Other | Other codes outside above categories | Other malignancies or unspecified sites |

**S2 Table.**
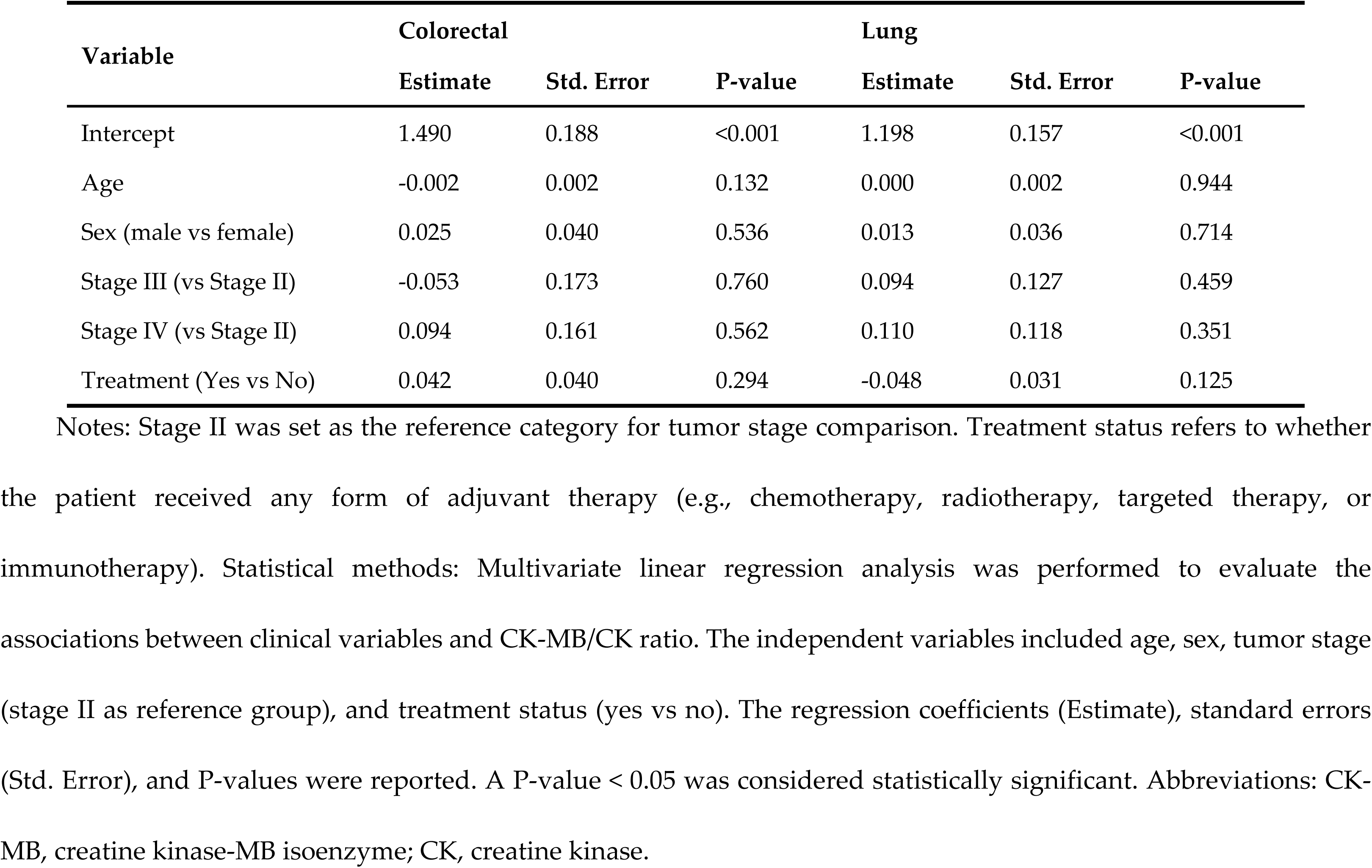
Multivariate linear regression analysis of factors associated with CK-MB/CK ratio in colorectal and lung cancer patients.

| Variable | Colorectal |  |  | Lung |  |  |
| --- | --- | --- | --- | --- | --- | --- |
|  | Estimate | Std. Error | P-value | Estimate | Std. Error | P-value |
| Intercept | 1.490 | 0.188 | <0.001 | 1.198 | 0.157 | <0.001 |
| Age | -0.002 | 0.002 | 0.132 | 0.000 | 0.002 | 0.944 |
| Sex (male vs female) | 0.025 | 0.040 | 0.536 | 0.013 | 0.036 | 0.714 |
| Stage III (vs Stage II) | -0.053 | 0.173 | 0.760 | 0.094 | 0.127 | 0.459 |
| Stage IV (vs Stage II) | 0.094 | 0.161 | 0.562 | 0.110 | 0.118 | 0.351 |
| Treatment (Yes vs No) | 0.042 | 0.040 | 0.294 | -0.048 | 0.031 | 0.125 |

Notes: Stage II was set as the reference category for tumor stage comparison. Treatment status refers to whether the patient received any form of adjuvant therapy (e.g., chemotherapy, radiotherapy, targeted therapy, or immunotherapy). Statistical methods: Multivariate linear regression analysis was performed to evaluate the associations between clinical variables and CK-MB/CK ratio. The independent variables included age, sex, tumor stage (stage II as reference group), and treatment status (yes vs no). The regression coefficients (Estimate), standard errors (Std. Error), and P-values were reported. A P-value < 0.05 was considered statistically significant. Abbreviations: CK-MB, creatine kinase-MB isoenzyme; CK, creatine kinase.

**S1 Fig.**
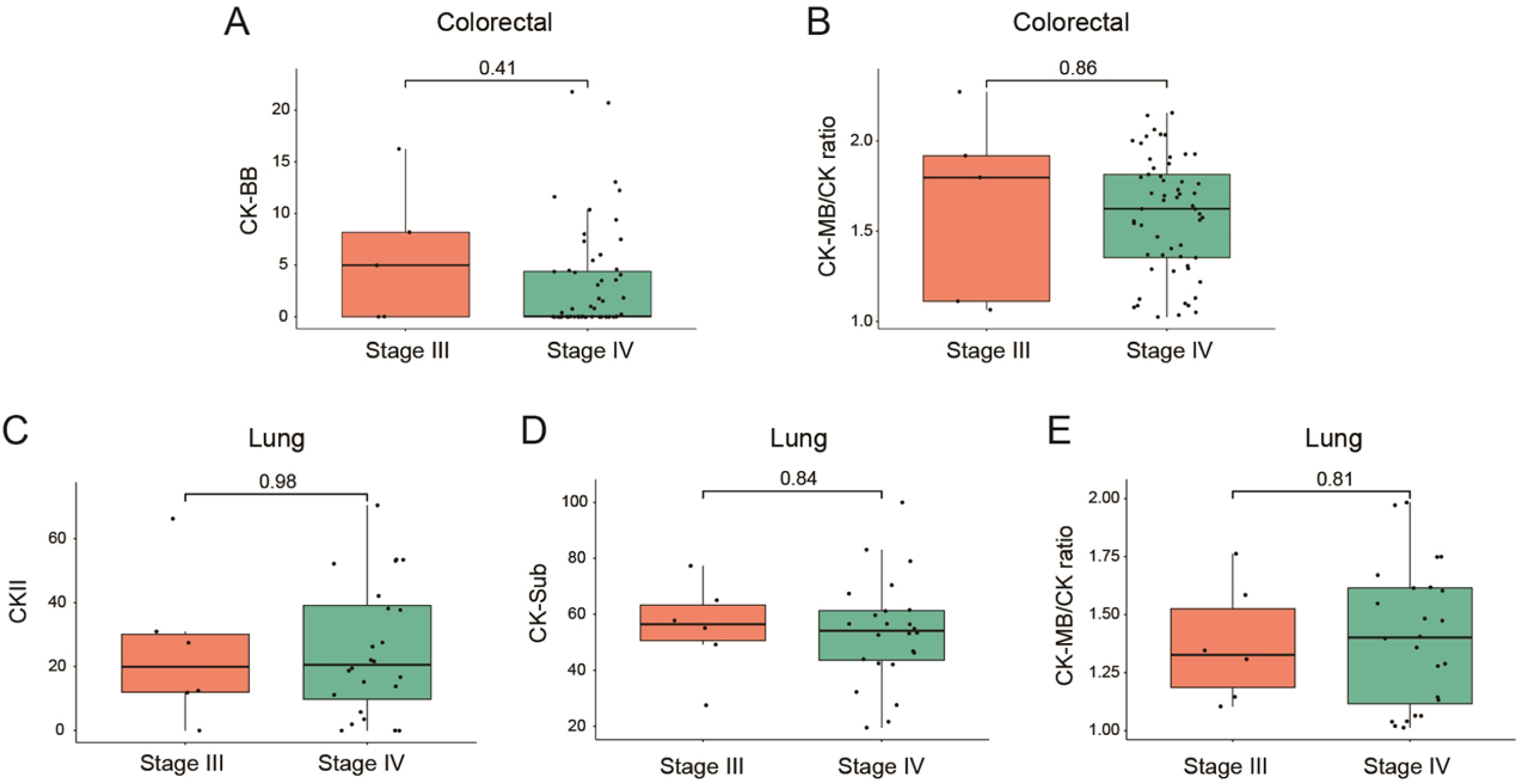
**Expression of serum CK isoenzymes in different disease stages**. (**A–B**) Comparison of CK-BB, CK-MB/CK ratio between stage III and stage IV colorectal cancer patients. (**C–E**) Comparison of CKII, CK-Sub, and CK-MB/CK ratio between stage III and stage IV lung cancer patients. Statistical methods: Group comparisons were performed using the Wilcoxon rank-sum test. Abbreviations: CK-MB, creatine kinase-MB isoenzyme; CK, creatine kinase; CKII, mitochondrial creatine kinase isoenzyme II; CK-BB, creatine kinase brain-type isoenzyme; CK-Sub, total CK isoenzyme activity (CK-BB + CKII + CKI).

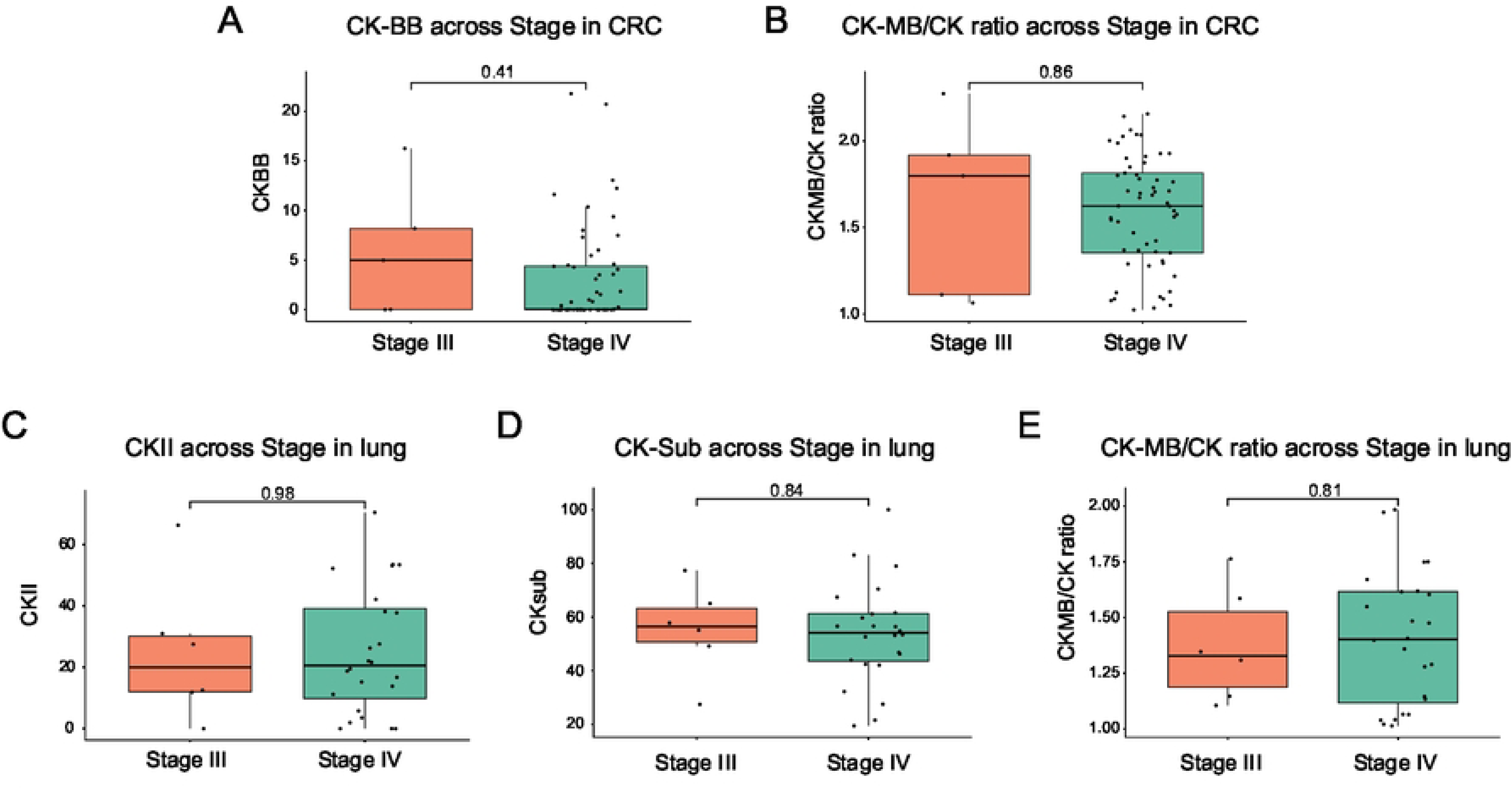

